# Safety and Immunogenicity of Fractional Doses of COVID-19 Vaccines among Nigerian Adults- A Randomised Non-Inferiority trial

**DOI:** 10.1101/2024.11.21.24317533

**Authors:** Salako Abideen Olurotimi, Musa Adesola Zaidat, Ige Fehintola Anthonia, Adam Abdullahi, Ayorinde Babatunde James, Ekama Sabdat, Odubela Oluwatosin, Idigbe Eugenia Ifeoma, Ajibaye Olusola, Altaf Mazharul, Adeneye Kazeem, Akinsolu Folahanmi T, Olojo Isimeme Ifedola, Okwuraiwe Azuka, Egharevba Henry, Ekpenyong Magaret, Elemuwa Uchenna, Ezenyi Ifeoma, Bitrus Fraden, Odubela Olayemi Rofiah, Oba Abdulrasheed, Idris Ganiu Adigun, Yusuf Jimoh, Akande Ibukun Ruth, Nwaiwu Stephine Ogechi, Omale Ojoma Louisa, Oyewunmi Oluwatobiloba Dorcas, Agbabiaka Adedoyin, Eyinade Olajumoke A, Ogunwale Joy, Abdullah Garba, Bello Yahya, Musa Baba Maiyaki, Ezejiofor Ogochukwu, Ejiro A. Ben, Iwalokun Bamidele Abiodun, Leah Rosenzweig, Obi Peter Adigwe, Adeyeye Christianah Mojisola, Faisal Shuaib, Wicek Witold, Yohhei Hamada, Ezechi Oliver Chukwujekwu, Ravindra K Gupta, Salako Babatunde Lawal

## Abstract

The shortage of COVID-19 vaccines posed a significant challenge in optimal response to the COVID-19 pandemic. Fractional doses of vaccine with adequate immunogenic response and proven safety profile emerged as potential strategy to extend the limited vaccine doses. This study was aimed to evaluate the immunogenicity and safety of fractional doses of the ChadOx1, Ad26.COV2.S, and BNT162B2 vaccines among healthy Nigerian adults. A non-inferiority multi-site triple-blind clinical trial was undertaken in Nigeria. Healthy Nigerian adults (18-65 years) who met the inclusion criteria were enrolled in the study. Participants were block-randomized into three vaccine arms (ChadOx1 quarter, half, and full dose; Ad26.COV2.S: quarter, half and full dose; and BNT162B2: half and full dose) . Participants, clinical staff (clinicians and nurses) and laboratory personnel were blinded. The primary objective of the study was to evaluate non-inferiority in seroconversion rates, defined as geometric mean fold rise (GMFR) ≥ 2.5 in serum anti-spike IgG titre at 28 days post-vaccination by ELISA. Immunogenicity analysis included use of serum neutralization assays using pseudotyped virus bearing spike from Wu-1 and Omicron variants. A total of 1891 participants were enrolled between June 21, 2022, and January 25, 2023. 320 participants in the fractional dose group and 220 in the standard dose group completed follow-up and were included in the analysis. SARS-CoV-2 seropositivity at baseline was high, at 68% (365/539). Seroconversion (geometric mean fold rise) was comparable between standard and fractional doses. For ChAdOx1, 31% achieved ≥ 2.5 fold change increase in serum binding antibody in the standard dose arm (16/52), 28% in half dose (15/53), and 34% in quarter dose (18/53). For Ad26.COV2.S, the proportions were 27% (28/105) in standard dose, 32% (22/68) in half dose, and 30% (21/71) in quarter dose arms respectively. For BNT162N2, the proportions were 43% (27/63) in standard dose and 39% (29/75) in half dose. Subset analysis of binding and neutralization responses in (n=64) participants demonstrated high degree of of prior exposure to SARS-CoV-2 ancestral and Omicron lineage variants prior to vaccination. Serum neutralization responses showed ≥2-fold response to both full and fractional doses indicating immunogenic responses to the vaccine dosing regimens. There was no report of serious adverse events. Fractional vaccine doses showed potential to generate non-inferior immune responses compared to standard doses in the context of a population with high rate of previous exposure to SARS-CoV-2 infection. The three vaccines are safe and well tolerated. Fractional dose should be considered to boost herd immunity and prevent outbreaks of SARS-COV-2.

## Introduction

The Coronavirus disease 2019 (COVID-19) pandemic, which began in December 2019, rapidly spread globally and resulted in over 700 million reported cases and approximately 7 million deaths.^1^ To mitigate the impact, multiple strategies, including infection control, social distancing, lockdown, and vaccination were introduced, each yielding different outcomes.^2–5^ Among these measures, vaccination was identified as the most effective public health intervention to prevent severe disease.^6,7^ The first COVID-19 vaccine was administered on December 8, 2020^8^ and to promote vaccine equity, the COVID-19 Vaccines Global Access (COVAX) Facility and the World Health Organization established distribution targets.^1^ However, due to vaccine shortfalls and logistical challenges, targets were not met by the end of 2021^9^ leading to uneven distribution of vaccines.^10–12^

Limited access to vaccines across low- and middle-income countries (LMICs) has limited the potential impact and benefit of vaccination in these settings, emphasizing the need for global vaccine equity and coverage.^9^ Data from 2021 showed vaccination rates across high-resources countries was 10-fold higher than in in the low-income countries, with Africa trailing significantly.^13^ Strategies to extend the use of limited vaccines have previously been explored and would have been beneficial if implemented early in the pandemic. One such strategy is the use of fractional vaccine doses, which involves administering reduced doses of vaccine to achieve adequate immune protection. This approach enables broader vaccine coverage within constrained resources and is associated with minimal side effects.^14–16^ The WHO has previously recommended dose fractionating for previous outbreaks, yielding favorable immunization outcomes and aiding epidemic control.^17^ Study evidence from Yellow Fever fractionated studies has shown similar antibody response outcomes relative to full doses with sustained long-term protection in vaccinated individuals.^14^

In the context of COVID-19 vaccination, studies from high-income countries show that fractional doses of mRNA vaccines could provide a robust immune response against COVID-19 ^15,18^. These studies however, often involved small sampling (<15) and were non-randomized. In sub-Saharan Africa, there are compelling reasons to determine the effectiveness of fractional doses of the COVID-19 vaccines, particularly due to limited capacity to secure adequate vaccine supplies for their large population. Nigeria with a population of over 200 million, would benefit significantly from fractional dosing strategies of the COVID-19 vaccines and provide critically lacking data from the sub-Saharan African region.

This study evaluated the safety and immunogenicity of fractional doses of the ChadOx1, Ad26.COV2.S, and BNT162B2 vaccines among healthy Nigerian adults. The primary objective of the study was to evaluate non-inferiority in seroconversion rates, defined as geometric mean fold rise (GMFR) ≥ 2.5 in serum anti-spike IgG titre at 28 days post-vaccination. Immunogenicity analysis included use of serum neutralization assays.

### Methodology Study Design

We conducted a multi-site, randomized, triple-blind, non-inferiority trial across five sites in five of Nigeria’s six geopolitical zones. The study sites were Aminu Kano University Teaching Hospital, Kano (Northwest), National Institute for Pharmaceutical Research and Development (NIPRD), Abuja, Federal Capital Territory (North Central), Nnamdi Azikiwe University Teaching Hospital, Anambra (NAUTH) (South-East), Delta State University Teaching Hospital (DELSUTH) Delta, (South-South) and the Nigerian Institute of Medical Research, Lagos (Southwest).

### Study population

Participants eligible for recruitment were adults (18 – 65 years). Exclusion criteria were: (i) previous SARS COV-2 infection defined by IgM anti-N positivity ii) pregnancy and breastfeeding iii) debilitating disease conditions or severe allergic reactions iv) previous COVID-19 vaccination v) current treatment with an investigational agent for prophylactic COVID-19 vi) individuals who were likely to travel during the study period. A total of 2491 communities across five trial sites (Kano, Abuja, Anambra, Delta, and Lagos states) in Nigeria (Figure 1A). Following screening, 1894 study participants were eligible and assigned to the vaccine arms. All enrolled participants had health insurance coverage for the period of the study. Participants were also required to provide plasma sample at five different timepoints during the study period. ChadOx1 and BNT162B2 vaccines had similar dosing schedules (prime and booster doses) for all study participants. Ad26.COV2.S recommended as a single dose regime and with more extended dosing timeline had a different schedule; full dose participants were not administered with a booster dose, other participants in the Ad26.COV2.S fractional dosing group had booster doses administered.

**Figure 1.**
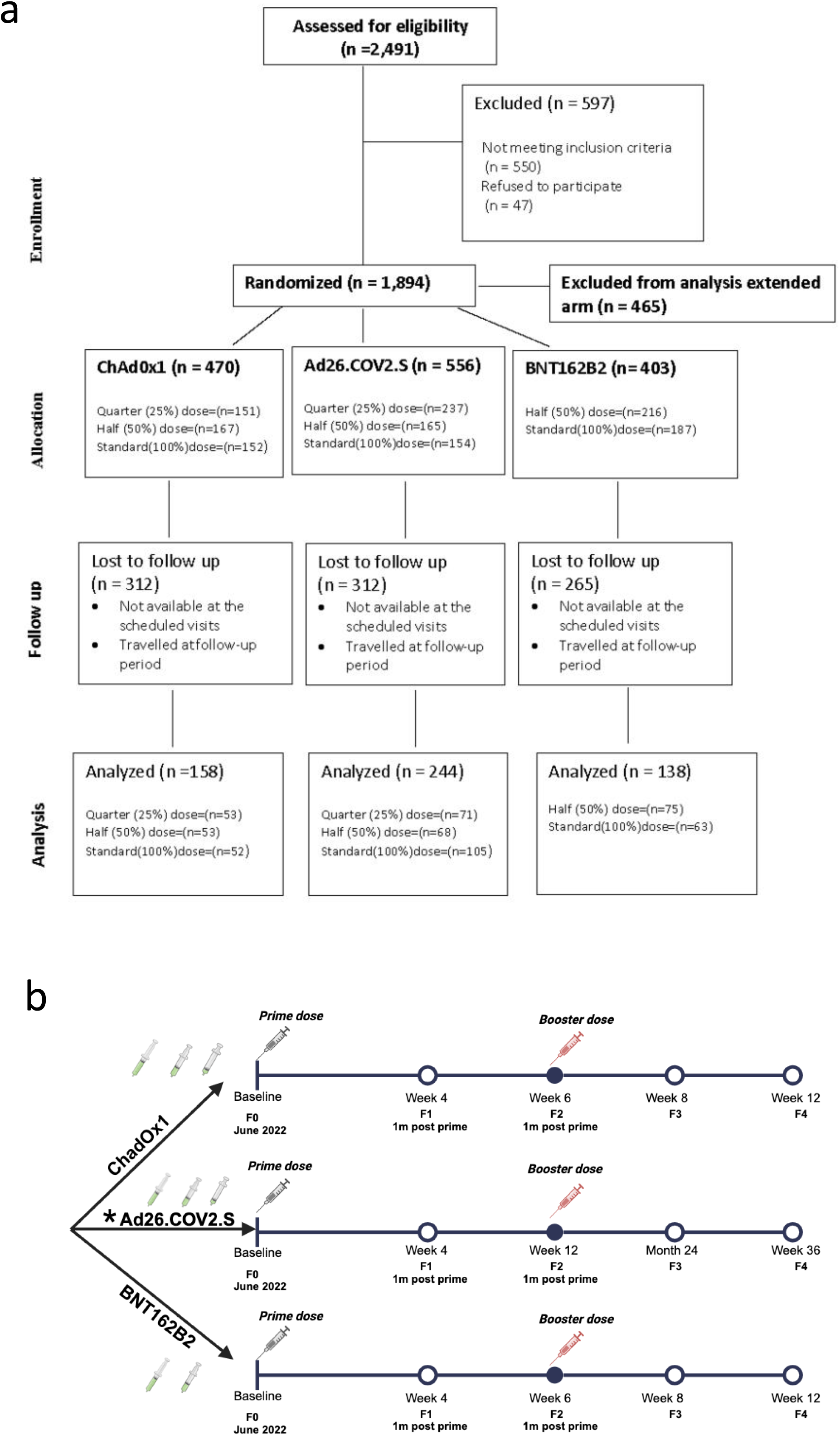
Study flow chart and disposition of patient recruitment . **a)** CONSORT diagram of the enrolment and screening, randomisation, and follow-up of participants randomized to receive full or fractionated doses of ChadOx1, Ad26.COV2.S and BNT162B2 vaccines. **b)** Disposition of study population showing timepoints of vaccination and study visits. * Participants randomized to receive full doses of AD26.COV2.S only received one dose as per standard recommendation and those randomized to half and quarter doses received booster doses at 12 weeks.

### Randomization and Blinding

The randomization was done by the lead statistician at the coordinating centre. Block randomization was used to allocate participants to different arms in blocks with each block containing a predetermined number of participants. The randomization sequence was concealed from the clinical staff (clinician and nurses), participants, and laboratory personnel to maintain blinding. This was done using sequentially numbered, opaque, sealed envelopes (SNOSE). The SNOSE was handed over to the study pharmacist in each site at initiation.

In each of the sites, each eligible study participant was allocated a unique enrolment identifier following a phone call by the site PI who communicates with the lead statistician. After clinical evaluation, the study participant was directed to the study pharmacist to decode and dispense the vaccine for the study nurse to administer.

### Follow-up

#### Safety-Monitoring

Safety data were reviewed at periodic intervals by the study safety review team and the data safety monitoring board. All enrolled participants were followed up with daily phone calls to ascertain any adverse event or complaint noticed within the first 72 hours of vaccine administration by the study nurse at each of the study sites. Clinical evaluation were done at baseline and follow-up visits at days 28, 42, 56, and 84 for ChadOx1 and Ad26.COV2.S vaccines (Days 28, 84, 180, and 252 for BNT162B2 vaccine); Figure 1B.

#### Immunologic evaluation

Blood samples were collected at baseline and follow-up visits. The study participants were withdrawn without replacement during the visit if they were pregnant or voluntary withdrawal with appropriate documentation.

### Study Vaccines and Dosing

Three COVID-19 vaccines (ChadOx1, Ad26.COV2.S, and BNT162B2 COVID-19 vaccines) were used for the trial (Table 1). The vaccines were supplied and distributed to all the study sites by the National Primary Health Care Development Agency (NPHCDA) who obtained the vaccines from the stock allocated for use in the country. Lot sampling, content verification, purity testing, and analysis were performed on the assigned vaccine batches for use in the study by the National Agency for Food Drug Administration and Control (NAFDAC). The first dose of the vaccines were administered at baseline (Day 0) and the second dose of the vaccines was administered on Day 42 (for ChadOx1, BNT162B2 vaccines) and Day 84 (for the Ad26.COV2.S vaccine).

**Table 1.** Baseline Characteristics of the Study Population based on the Primary Vaccine dose (n= 540)

|  | Vaccine type |  |  |
| --- | --- | --- | --- |
| Variables | ChadOx1<br>(n=158) | BNT162B2<br>(n=138) | AD26.COV2.S<br>(n=244) |
| **Mean age (± SD) years** | 36.3 ± 12.8 | 36.4 ± 13.0 | 37.1 ± 13.6 |
| **Age Group n(%)** |  |  |  |
| 18-55 years | 147(93.0) | 125(90.6) | 218(89.3) |
| 56-65 years | 11(7.0) | 13(9.4) | 26(10.7) |
| **Sex** |  |  |  |
| Female | 79(50) | 82(59.4) | 161 (66) |
| Male | 79(50) | 56(40.6) | 83(34) |
| **Marital Status** |  |  |  |
| Married | 84(53.2) | 72(52.2) | 135(55.3) |
| Unmarried | 74(46.8) | 66(47.8) | 109 (44.7) |
| **Educational Level** |  |  |  |
| None | 10 (6.3) | 5(3.6) | 20 (8.2) |
| Primary | 20 (12.7) | 29(21) | 41(16.8) |
| Secondary | 81(51.3) | 67(48.6) | 126(51.6) |
| Tertiary | 47 (29.7) | 37(26.8) | 57(23.4) |
| **Occupation** |  |  |  |
| Employed | 116(73.4) | 107(77.5) | 194 (79.5) |
| Unemployed | 42(26.6) | 31(22.5) | 50(20.5) |
| **Average Income** |  |  |  |
| 1-30,000 | 67(42.4) | 62(44.9) | 136(55.7) |
| Above 30,000 | 34(21.5) | 25(18.1) | 46(18.9) |
| **Seropositivity baseline** |  |  |  |
| **Negative** | 56(35.4) | 57(41.3) | 61(25) |
| **Positive** | 101(63.9) | 81(58.7) | 183(75) |

### Procedure for vaccine storage, fractional doses preparation and administration

The Ad26.COV2.S and BNT162B2 COVID-19 vaccines have pre- and post-thawing temperature storage and shelf-life conditions. Ad26.COV2.S and BNT162B2 COVID-19 vaccines were originally stored at -90°C to -60°C and -25°C respectively at the national cold storage facility by the NPHCDA after which they were thawed and then distributed at 2 - 8°C to the various study sites. The BNT162B2 vaccine required reconstitution with a diluent (Sodium Chloride injection 0.9% USP) before use. The ChadOx1 vaccine was originally stored and distributed at 2 - 8°C. At study sites, temperature of the vaccine storage refrigerators were charted twice daily, and the recordings were documented. All the vaccines have a shelf life of 6 hours once a vial is opened, a new expiry time is calculated and documented and the left over from the multi-vials were discarded after 6 hours.

Standardized and calibrated syringes of 0.3 mL, 0.5 mL and 1 mL capacity were used to withdraw and prepare the prefilled full and fractional doses of the vaccines as required. All prefilled and reconstituted vaccines were kept at 2 - 8°C in vaccine carriers with thermometers until ready for use. The model employed in this study for vaccine administration was such that the trial pharmacists prepared a pre-filled syringe of the full or fractional doses (quarter (25%) or half (50%) doses) just before administration and then handed over to the study nurse for administration to the trial participants. The vaccines were administered intramuscularly in the upper arm in the deltoid muscle with either 0.3 mL, 0.5 mL or 1 mL auto-disable syringes (detachable needle size 23G x 1 ¼’) at 90° injection angle.

### Sample Transportation

Blood samples were collected at the different sites. The samples for serological analysis were duly labelled and temporarily stored in 0.5 mL aliquots at -20° C freezers. The samples were transported (airfreight) on dry ice in insulated Styrofoam packs from the sentinel laboratories to the SIFCoVAN Central Laboratory at NIMR, Lagos within 4 weeks of sample collection. The duration of sample transportation were about 60-120 minutes. However, complete blood count, electrolyte, urea, creatinine, liver function test and urinalysis were done at the enrolment sites based on routine clinical care and best practices. The generated results from the assay were entered into the case report form and then into the study database (web-based version of RedCap^R^).

### Anti-SARSCoV-2 Spike protein IgG antibody Testing by ELISA

A solid-phase sandwich Enzyme-linked immunosorbent assay (ELISA) was performed to determine the concentrations of anti-SARS-CoV-2 IgG in the serum samples of each of the healthy volunteers vaccinated with standard or fractional doses of the tested COVID-19 vaccines at baseline (day 0) and the different time points in the study. After the collection of whole blood (5 mL) into a plain tube, serum was recovered by centrifugation at 3000 rpm for 10 min at room temperature and transferred to a new pre-labelled plain tube. Ten microlitres of the recovered serum were used immediately or stored at -20°C in preparation for ELISA screening. The ELISA assay detects IgG antibodies directed against the viral spike protein receptor-binding domain (S-RBD) and 0.89 U/ml was used as the cut-off value for qualitative determination of antibody positivity at baseline. For the quantitative assay, each serum sample was pre-diluted 100-fold (1: 99 v/v) in assay buffer and used for analysis according to the manufacturer’s protocol (Thermo Fisher Scientific, Vienna Austria) using the Human SARS-CoV-2 Spike (Trimer) IgG sandwich ELISA kit (Catalog Number BMS2325, Lot Number 359554016).

### Binding antibody measurements to S and N by Luminex assay

In a subset of participants with paired available sampling across baseline(F0), Day 28 or week 4 (F1) and Week 8 or 24 (F3) time points (n=64), we assessed binding antibodies using the highly sensitive Luminex assay. We measured binding antibodies against SARS-COV-2 trimeric spike protein (S), nucleocapsid protein (N), Wu-1 D614G and Omicron (BA.1) specific receptor-binding domain (RBD) as previously described which were validated using pre-pandemic samples.^19^

### Neutralizing antibody measurement by lentiviral pseudotype neutralization assay

Neutralizing antibodies were measured using the SARS-CoV-2 lentiviral pseudovirus (PV) technique; this was prepared by transfecting HEK293T cells with Wu-1-614G wild type (WT), BA.1, BA.2 and XBB plasmids in conjunction with p8.91 HIV-1 gag-pol expression vectors as previously described.^20,21^ There is concordant evidence to indicate a high degree of correlation between PV and live virus neutralisation.^22,23^ Briefly, plasma samples were inactivated at 54 °C for 60 minutes, serially diluted in duplicates and incubated with PVs at 37 °C for another 60 minutes preceding the addition of Hela-ACE2 cells. The plasma dilution/virus mix was incubated for 48 hours in a 5% CO2 incubator at 37 °C, and a luminescence reading was measured using the Bright-Glo Luciferase assay system (Promega).^20^ Neutralization was calculated relative to virus-only controls after a cell normalization protocol as a mean neutralization with s.e.m, half maximum inhibitory dose (ID50). This was calculated via the GraphPad Prism software version 9.3.1 and ID50 > 20 units was considered positive.

### Statistical analysis

Statistical analysis was performed to assess the safety and immunogenicity of fractional doses of the ChAdOx1, AD26.COV2.S, and BNT162B2 COVID-19 vaccines.

Sample size calculation: The trial sample size was calculated based on several key factors: a power of 80% (1-β), a significance level of 5% (α), an expected seroconversion rate of 92% in the control group (100% dosing arm), and 80% in the experimental arm. A non-inferiority margin was set at 20% (d) and we considered an estimated attrition rate of 10%. A total sample size of 1812 participants were calculated with 151 participants per arm. Figure 1A. Safety data were analyzed by comparing the incidence of adverse events (AEs) among participants receiving fractional doses with those receiving full doses, using descriptive statistics and chi-squared tests.

Immunogenicity was evaluated by measuring the geometric mean concentration (GMCs) of SARS-COV2 antibodies at baseline and at 28 days post-vaccination time point, and their ratios were estimated. Next, we estimated the percentage of participants who achieved seroconversion, along with 95% confidence intervals (CIs), using the exact Clopper-Pearson method by vaccine dose and type. Seroconversion was defined as a Geometric Mean Fold Rise (GMFR) of ≥ 2.5. To assess the non-inferiority of the seroconversion rates, we estimated the point difference between the fractional and standard dose arms with 95% CIs using the Wilson score method. Fractional doses were considered non-inferior if the lower bound of the confidence interval (CI) for the difference in seroconversion exceeded 20% (Figure 2A). These intervals were then transformed to express the ratio of GMC and GMF for the fractional dose relative to the standard dose. Immunogenicity outcomes were evaluated among participants who had paired visits at baseline and week 28. Both an overall analysis and a sub-analysis of seronegative participants were performed. All statistical analyses were conducted using STATA software, with a significance level set at p < 0.05 for determining statistical significance

**Figure 2.**
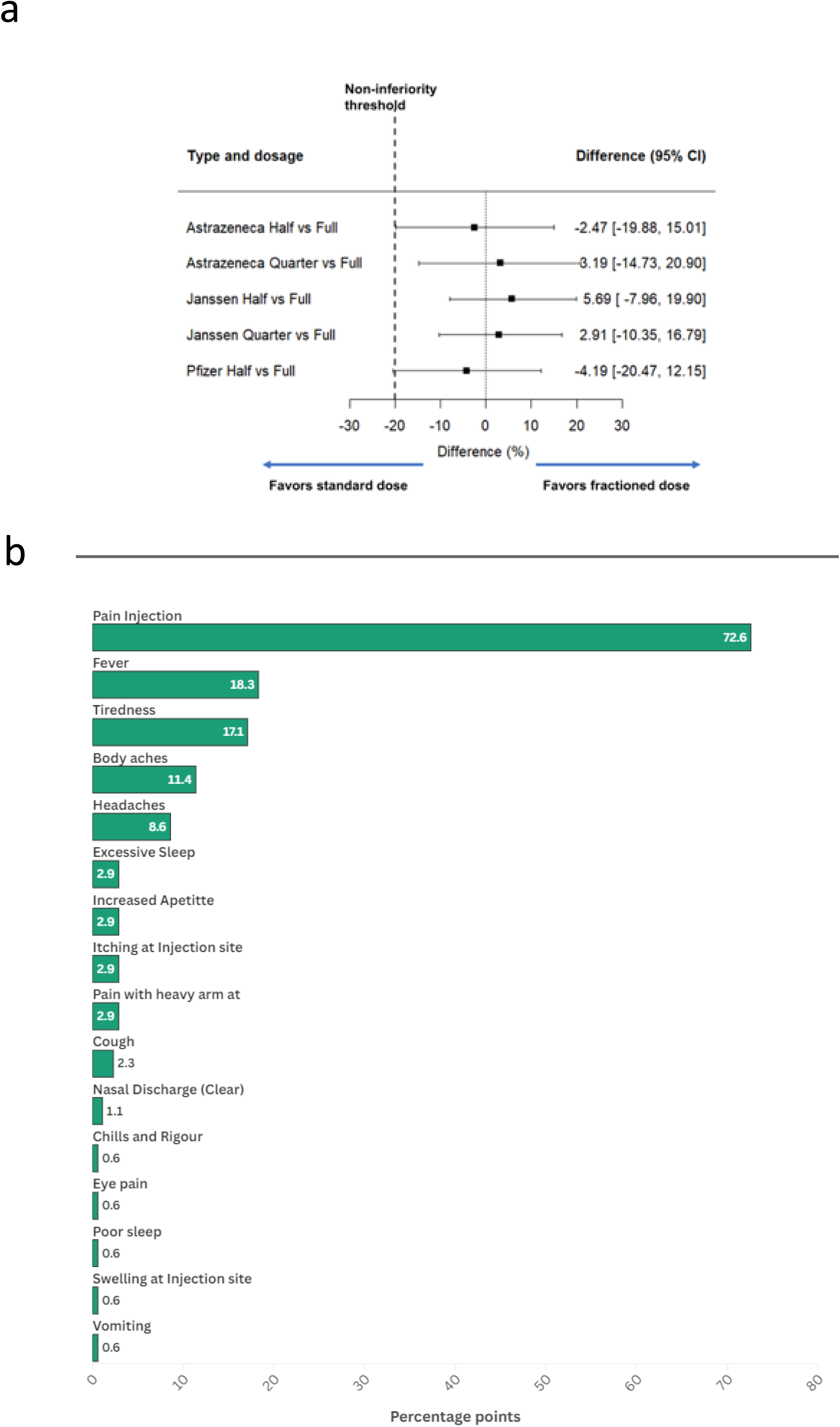
Primary outcome analysis and adverse events among the aggregated study participants (n=540) **a)** proportion of participants across study arms with geometric mean fold rise (GMFR) ≥ 2.5 in serum anti-spike IgG titre at 28 days post-vaccination. Non-inferiority margin was set at 20% and point difference between the fractional and standard dose arms with 95% CIs using the Wilson score method. **b)** proportion of study participants reported at least one adverse event occurring within the 72 hours following COVID-19 vaccine administration, irrespective of the study arm

### Ethics

The study protocol was approved by the National Health Research Ethics Committee of Nigeria (NHREC/01/01/2007-28/11/2021) and approval for the conduct of the clinical trial in the country was granted by the National Agency for Food and Drug Administration and Control (NAFDAC/DER/VCTD/SIFCoVAN/2022). In addition, all the trial sites obtained ethical approval from their respective local ethics committee [Institutional Review Board (IRB), Nigerian Institute of Medical Research (IRB-22-016), Ministry of Health, Kano State (NHREC/17/03/2018), Health Research Ethics Committee, Delta State University Teaching Hospital (HREC/PAN/2022/021/0469), NIPRD Health Research Ethics Committee (NHREC/039/21A), and NAUTH Health Research Ethics Committee (NAUTH/CS/66/VOL.15/VER.3/053/2022/036)]. According to the Declaration of Helsinki, the trial complied with good clinical practice guidelines. The protocol was registered with the Pan African Clinical Trials Registry (PACTR) with approval number PACTR 202206754734018.

## Results

A total of 2491 study participants were screened between June 21, 2022, and January 25, 2023. 1894 study participants were eligible and assigned to the vaccine arms (ChadOx1 n=611; Ad26.COV2.S n=706; BNT162B2 n=577, Supplementary Table 1). Study participants (n= 465) on the extended vaccine dose were excluded from this analysis bringing sample size to 1429. Included in the analysis for ChadOx1 arm, were 152, 151 and 167 participants for the standard, quarter and half fractional doses respectively. For the Ad26.COV2.S arm, 154, 237 and 165 study participants were allotted to the standard, quarter and half fractional doses respectively. For the BNT162B2 vaccine arm, 187 and 216 study participants were assigned to the standard and half fractional dose respectively. The attrition rate was 59% by the day 28 visit.

The primary outcome analysis per protocol included 320 participants in the fractional dose group and 220 in the standard dose group (Supplementary Table 2). The mean age (SD) of the participants was 36.7 (13.2) years with a female-to-male distribution of 1.5:1. The mean age (SD) of the participants in the quarter, half, and full dose arms (irrespective of the vaccine type) were 34.0 (± 13.1), 33.9 (± 12.9) and 35.1(± 12.9) years respectively. Three hundred and sixty-five out of five hundred and forty (67.6%) participants had a positive SARS-COV-2 antibody at baseline. The proportion of participants with positive SARS-COV-2 antibodies at baseline was found to be approximately equally distributed across the arms-quarter dose (69.4%), half dose (66.2%) and full dose (68.2%). (p = 0.822) (Table 1).

At 28 days post-vaccination, 177 out of 540 participants (21.7%) showed a rise in GMFR a 2.5-fold rise in anti-SARS-COV-2 titre from baseline irrespective of the vaccine arm or dose. A total of 72 out of 220 in the full dose (32.7%) and 105 of 320 (32.8%) in the fractional doses (quarter and half dose) irrespective of the vaccine arm seroconverted at day 28 (Table 2).

**Table 2.** Increases in SARS-CoV-2 Spike specific binding IgG in the study population per arm and vaccine type following one dose.

| | n/N | % (95% CI)<br>With serum<br>IgG ≥ 2.5 | Difference | Risk ratio | Geometric Mean<br>Concentration<br>(95% CI) | GMR |
| --- | --- | --- | --- | --- | --- | --- |
| **ChAdOx1** |  |  |  |  |  |  |
| Standard Dose | 16/52 | 30.8 (18.7-45.1) | - |  | 1.34(0.91-1.96) |  |
| Half (50%) Dose | 15/53 | 28.3 (16.8-42.3) | -2.5 (-19.9-15) | 0.89 (0.38-2.06) | 1.45(0.99-2.11) | 1.08 |
| Quarter (25%) Dose | 18/53 | 34 (21.5-48.3) | 3.2 (-14.7-20.9) | 1.16 (0.51-2.64) | 1.81(1.13-2.91) | 1.08 |
| **Ad26.COV2.S** |  |  |  |  |  |  |
| Standard Dose | 28/105 | 26.7 (18.5-36.2) | - |  | 1.51(1.13-2.03) |  |
| Half (50%) Dose | 22/68 | 32.4 (21.5-44.8) | 5.7 (-8-19.9) | 1.32 (0.67-2.56) | 1.72(1.20-2.46) | 1.13 |
| Quarter (25%) Dose | 21/71 | 29.6 (19.3-41.6) | 2.9 (-10.3-16.8) | 1.15 (0.59-2.25) | 1.19(0.85-1.64) | 0.78 |
| **BNT162B2** |  |  |  |  |  |  |
| Standard Dose | 27/63 | 42.9 (30.5-56) | - |  | 2.68 (1.90-10.12) |  |
| Half (50%) Dose | 29/75 | 38.7 (27.6-50.6) | -4.2 (-20.5-12.1) | 0.84 (0.42-1.66) | 1.99(1.31-2.99) | 0.73 |

In the ChadOx1 vaccines arm, 30.7%, 28.3% and 33% of the participants in the full, half and quarter doses of the vaccine achieved seroconversion. The difference in seroconversion rates were -2.5% (95% CI: -19.9 to 15.0) for the half dose vs. full dose, and 3.2% (95% CI: -14.7 to 20.9) for the quarter vs full dose, with both fractional doses meeting the non-inferiority criteria. Among study participants in the Ad26.COV2.S vaccines, 26.7%, 30.9%, and 29.6% in the full, half and quarter doses of the vaccine achieved seroconversion. The differences in the seroconversion were 5.7% (95% CI: -8.0 to 19.9) for the half dose vs. full dose and 2.9% (95% CI: -10.3 to 16.8) for the quarter vs. full dose; with both fractional doses considered non-inferior to the full dose. In the BNT162B2 vaccine arm, 44.4% and 40% of the participants in the full and half dose had a rise in GMFR 2.5-fold-rise (Table 2). The half dose (BNT162B2 vaccine arm) very marginally failed to meet the non-inferiority criteria with a difference of -4.2% (95% CI: -20.5 to 12.1) (Figure 2A; Table 2).

On sub-analysis of the participants with negative serostatus for anti-SARS-COV-2 at baseline, 112 of 177 (63.3%) seroconverted irrespective of the vaccine arm or dose. In the ChadOx1 vaccines arm, 68.8%, 80% and 66.7% of the participants in the full, half and quarter doses of the vaccine achieved seroconversion. The difference in seroconversion rates were -10.3% (95% CI: -40 to 23.4) for the half dose vs. full dose, and 6.4% (95% CI: -35.9 to 26.2) for the quarter vs full dose, with both fractional doses not meeting the non-inferiority criteria. Among the participants in the Ad26.COV2.S vaccine group, 60.7%, 61.9%, and 47.6% of the participants in the full, half and quarter doses of the vaccine achieved seroconversion. The differences in the seroconversion were 9.1% (95% CI: -19.7 to 36) for the half dose vs. full dose and 1.6% (95% CI: 29.2 to 30.9) for the quarter vs. full dose; with both fractional doses failing to reach non-inferiority compared to the full dose. In the BNT162B2 vaccine arm, 60% and 63.3% of the participants in the full and half dose seroconverted, though half dose did not meet the non-inferiority criteria with a difference of -3.7% (95% CI: -28.1 to 21.2; Figure 2A). In the sub-group analysis, there is a wide confidence interval due to the small number of participants (Table 3).

**Table 3.**
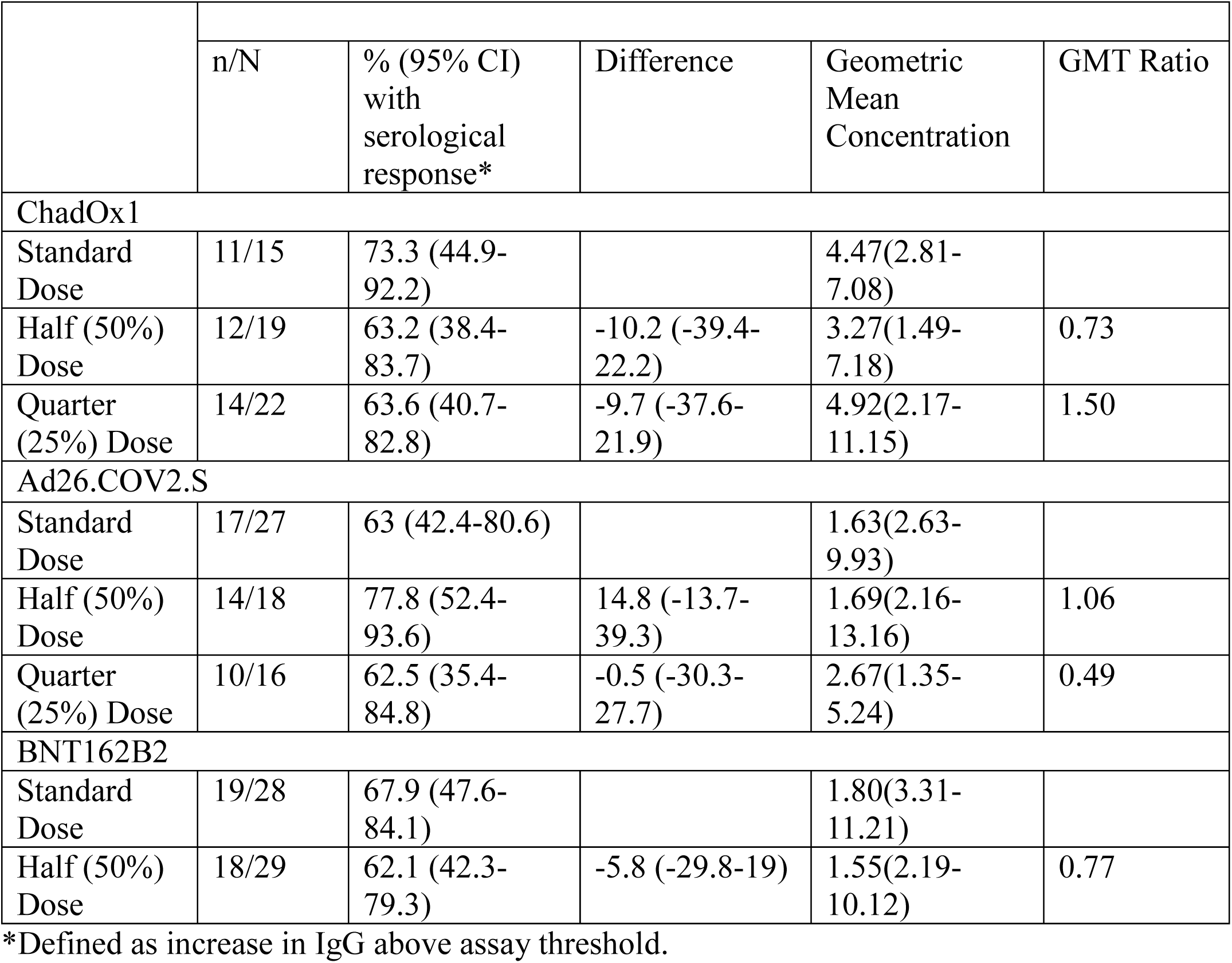
SARS-CoV-2 Spike specific binding IgG responses in the study population per arm and vaccine type following one dose in those seronegative at baseline.

|  | n/N | % (95% CI)<br>with<br>serological<br>response* | Difference | Geometric<br>Mean<br>Concentration | GMT Ratio |
| --- | --- | --- | --- | --- | --- |
| ChadOx1 |  |  |  |  |  |
| Standard Dose | 11/15 | 73.3 (44.9-92.2) |  | 4.47(2.81-7.08) |  |
| Half (50%) Dose | 12/19 | 63.2 (38.4-83.7) | -10.2 (-39.4-22.2) | 3.27(1.49-7.18) | 0.73 |
| Quarter (25%) Dose | 14/22 | 63.6 (40.7-82.8) | -9.7 (-37.6-21.9) | 4.92(2.17-11.15) | 1.50 |
| Ad26.COV2.S |  |  |  |  |  |
| Standard Dose | 17/27 | 63 (42.4-80.6) |  | 1.63(2.63-9.93) |  |
| Half (50%) Dose | 14/18 | 77.8 (52.4-93.6) | 14.8 (-13.7-39.3) | 1.69(2.16-13.16) | 1.06 |
| Quarter (25%) Dose | 10/16 | 62.5 (35.4-84.8) | -0.5 (-30.3-27.7) | 2.67(1.35-5.24) | 0.49 |
| BNT162B2 |  |  |  |  |  |
| Standard Dose | 19/28 | 67.9 (47.6-84.1) |  | 1.80(3.31-11.21) |  |
| Half (50%) Dose | 18/29 | 62.1 (42.3-79.3) | -5.8 (-29.8-19) | 1.55(2.19-10.12) | 0.77 |

\*Defined as increase in IgG above assay threshold.

A total of 929/1429 participants were evaluated for AE, based on phone call responses within 72 hours. A total of one hundred and seventy-five study participants reported at least one adverse event occurring within the 72 hours following COVID-19 vaccine administration, irrespective of the study arm. The common adverse events, irrespective of the vaccine arm, were pain at the injection site (72.6%), fever (18.3%), tiredness (17.1%), and body aches (11.4%), (Figure 2B). Most of the study participants did not report adverse events (Grade 0). However, of those with reported AE, most (79.4%) were of mild severity with no effect on daily activities (Grade 1), while 20.6% were of moderate severity associated with sleep disturbances and tiredness (Grade 2). There were no significant differences in adverse events by sex and age. There was no significant difference in the adverse events reported by participants in the fractional group compared to the standard dose group irrespective of the vaccine type (Supplementary Table 3).

We conducted a sub-analysis of binding antibody analysis in (n=64) participants with paired samples available at F0, F1 and F3 comprising (n=8) participants each across all eight vaccine groups. We first characterized at baseline, the proportion of these participants who were previously exposed to SARS COV-2 and SARS COV-2 Omicron variant defined by positivity to IgG anti-N and IgG anti-RBD Omicron using the Luminex assay. We observed 52/64 (81%) and 60/64 (94%) participants positive for IgG anti-N and IgG anti-RBD Omicron. We also observed 62/64 (97%), 61/64 (95%) and 62/64 (97%) for IgG anti-Spike (S), IgG Wu-1 anti-Receptor Binding Domain (Wu-1 RBD) and Wu-1 anti-Spike-1 (Wu-1 S1) respectively (Supplementary figure 1). We then characterized vaccine breakthrough infection in this subset of participants by assessing proportion of participants with a ≥2-fold increase in IgG anti-N during follow up time points relative to baseline or the preceding timepoint. We found that around a quarter of participants (17/64; 26%) experiencing a vaccine breakthrough infection.

We further characterized neutralizing antibody responses in (n=62) participants with sufficient samples available at F0, F1 and F3 for analysis. This comprised (n=8) participants from all vaccine groups, other than half-dose Ad26.COV2.S group which has (n=6) participants. In our neutralization analysis across the eight vaccine groups, the median pre-vaccination geometric mean neutralization titer against Wu-1 wild-type virus was 1473 (IQR 1242-2215). We observed similar rates of enhancement across the three vaccine types and doses following prime and booster doses (Figure 3-6). In the BNT162b2 group, against ancestral Wu-1 virus, prime dose vaccination increased neutralization antibody titers by 4.8 fold [GMT: 2390 to 11320] and 6.2-fold [GMT: 1487 to 9348] for full and half doses and the booster dose increased neutralization antibody titers by 1.7 fold [GMT: 11320 to 19930] and 2.1 fold [GMT: 9348 to 20320] respectively. In the ChadOx1 group, the prime dose increased neutralization antibody titers by 3.0 fold [GMT: 1223 to 3650], 4.1 folds [GMT: 1299 to 5310] and 3.7 folds [GMT: 763 to 2810] for full, half and quarter doses and the booster dose increased neutralization antibody titers 2.6 fold [GMT: 3650 to 9581], 2.1 fold [GMT: 5310 to 11384] and 5-fold [GMT: 2810 to 13746] respectively. In the Ad26.COV2.S group, the prime dose increased neutralization antibody titers by 3.6 fold [GMT: 3022 to 10912] 3.1 fold [GMT: 1458 to 4551] and 2.1 folds 3.6 fold [GMT: 1691 to 3642] for full, half and quarter doses and the booster dose increased neutralization antibody titers by 1.9 fold [GMT: 10912 to 20279], 2.5 fold [GMT: 4551 to 11160] and 3 fold [GMT: 3642 to 10880] respectively. Similar neutralization antibody titer enhancement was observed following all vaccinations tested (Figure 6A) suggesting that, in this largely exposed SARS-CoV-2 population, fractioning vaccine doses (half and quarter doses), elicits a similar breadth of responses relative to full doses. When comparing the impact of variants on antibody sensitivity to vaccine induced responses, we observed a consistent (<3 fold) reduction in sensitivity against BA.1 and BA.2 viruses relative to Wu-1 (Supplementary figure 2-4) despite recent evidence of exposure to Omicron variant, suggesting imprinted immunity.^24^ Notably, we also observed significant evasion of neutralizing antibody responses by XBB despite vaccination and natural infection (>10 fold across vaccine types and doses; Figure 6B).

**Figure 3:**
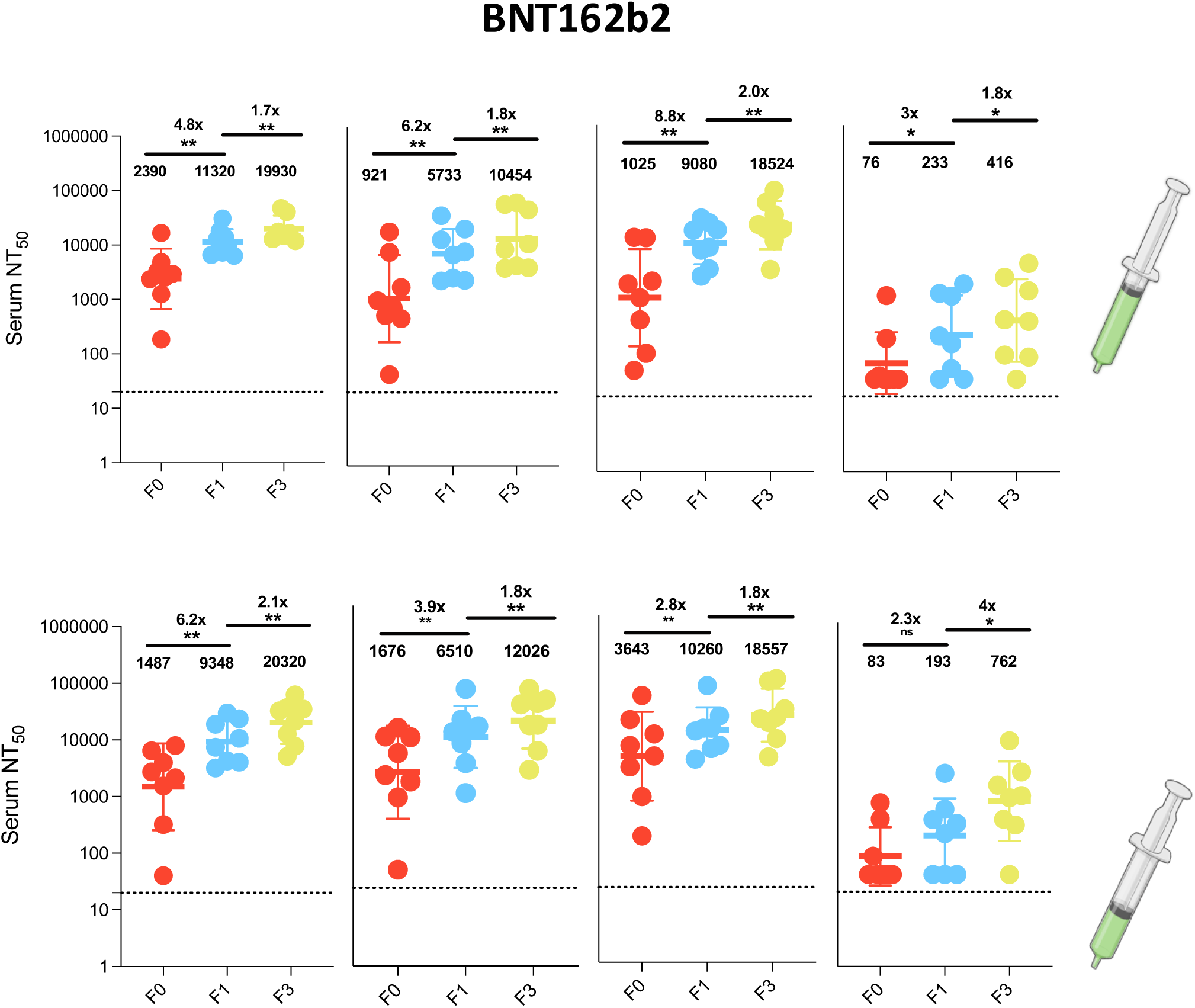
**Antibody responses to vaccination with BNT162b2 at full and half doses**. a): Plasma neutralization of pseudovirus against Wild type (Wu-1), BA.1, BA.2 and XBB after two doses of the BNT162b2 in Nigerian HIV-negative participants in Lagos Nigeria at three consecutive time points– baseline – F0 (before first-dose vaccination),F1 (1 month after 1st dose vaccination) and F3 (6 weeks post second dose at full doses (n=8) and half doses (n=8). Data points were compared using Wilcoxon test and shown as geometric mean titre (GMT) with 95% CI. \**P* < 0.05; \*\**P* < 0.01; \*\*\**P* < 0.001; \*\*\*\**P* < 0.0001; ns = not significant. Fold change are represented above the horizontal comparative lines.

**Figure 4:**
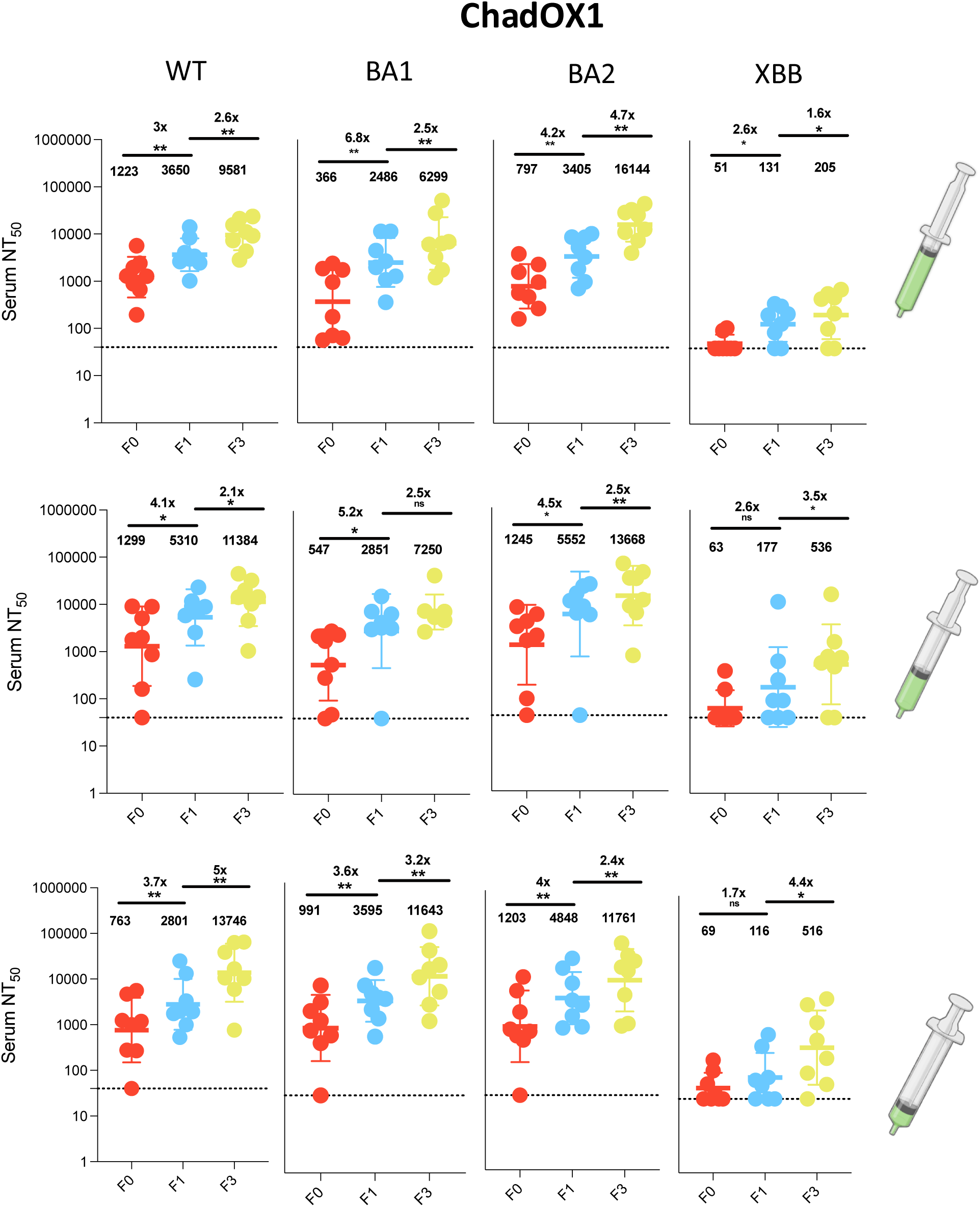
**Antibody responses to vaccination with ChadOX1 at full, half and quarter doses.** a): Plasma neutralization of pseudovirus against Wild type (Wu-1), BA.1, BA.2 and XBB after two doses of the ChadOX1 in Nigerian HIV-negative participants in Lagos Nigeria at three consecutive timepoints– baseline – F0 (before first-dose vaccination), F1 (1 month after 1st dose vaccination) and F3 (6 weeks post second dose at full, (n=8); half (n=8) and quarter doses (n=8). Data points were compared using Wilcoxon test and shown as geometric mean titre (GMT) with 95% CI. \**P* < 0.05; \*\**P* < 0.01; \*\*\**P* < 0.001; \*\*\*\**P* < 0.0001; ns = not significant. Fold change are represented above the horizontal comparative lines.

**Figure 5:**
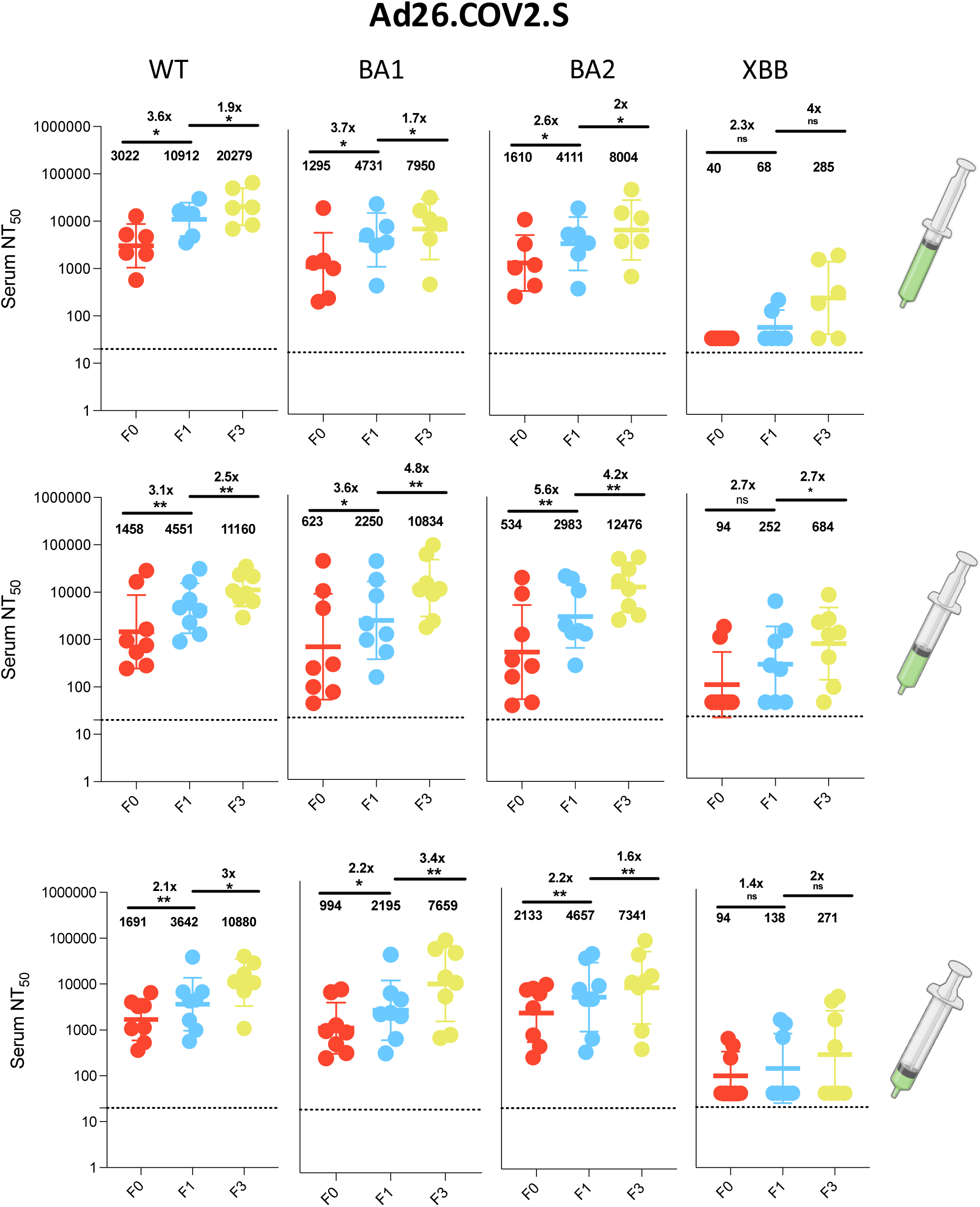
**Antibody responses to vaccination with Ad26.COV2.S at full, half and quarter doses.**a): Plasma neutralization of pseudovirus against Wild type (Wu-1), BA.1, BA.2 and XBB after two doses of the Ad26.COV2.S in Nigerian HIV-negative participants in Lagos Nigeria at three consecutive time points– baseline – F0 (before first-dose vaccination), F1 (1 month after 1st dose vaccination) and F3 (6-weeks post second dose at full, (n=6); half (n=8) and quarter doses (n=8). Data points were compared using Wilcoxon test and shown as geometric mean titre (GMT) with 95% CI. \**P* < 0.05; \*\**P* < 0.01; \*\*\**P* < 0.001; \*\*\*\**P* < 0.0001; ns = not significant. Fold change are represented above the horizontal comparative lines.

**Figure 6:**
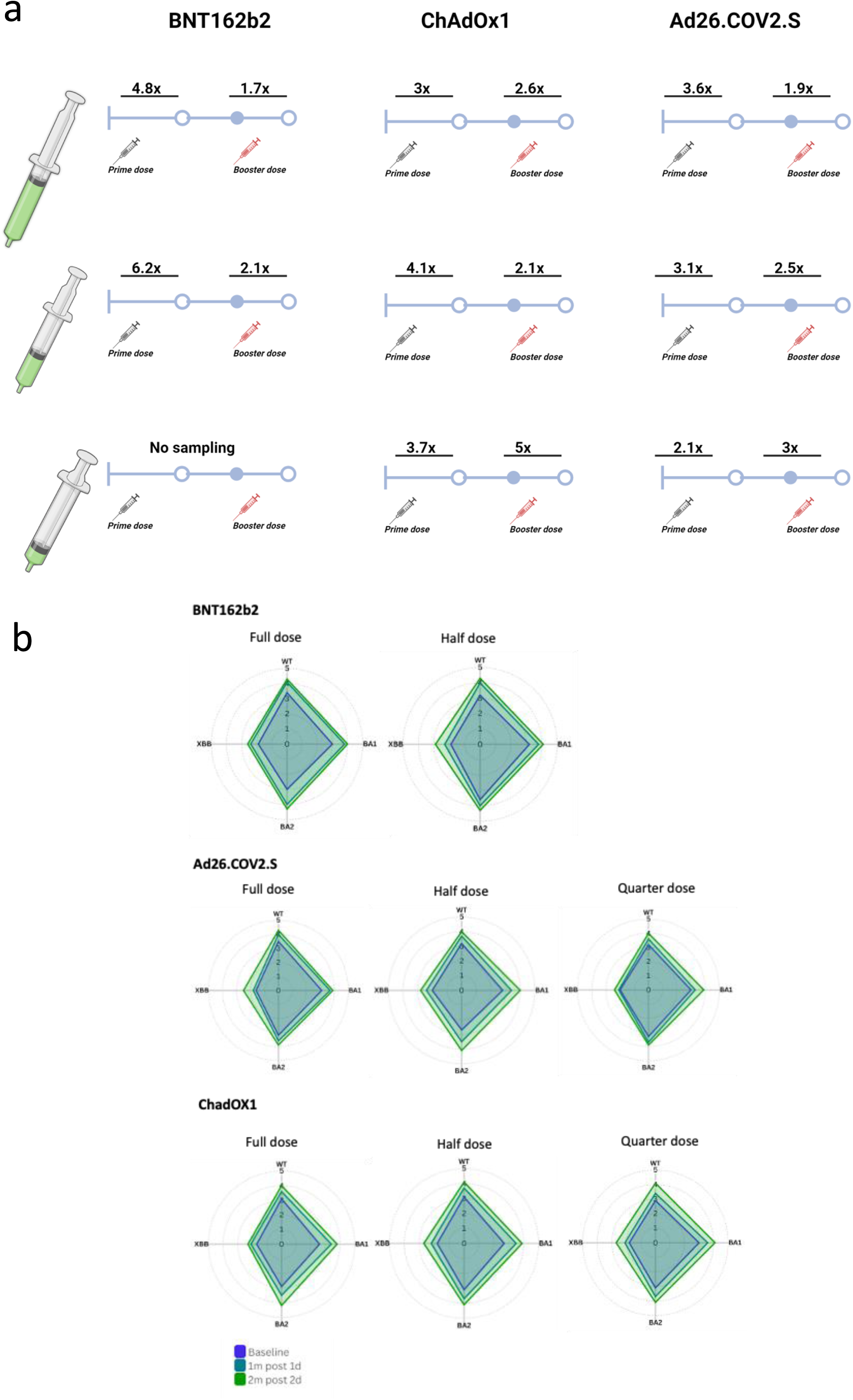
a). Fold-change following two doses of fractional doses of Ad26.COV2.S, ChadOX1 and BNT162b2. Plasma neutralization of pseudovirus against Wild type (Wu-1) fold-change following two doses of the Ad26.COV2.S, ChadOX1 and BNT162b2 doses either as full or quarter or half fractionated doses. Fold Change is shown above vaccine doses. **b)**. Antibody responses to vaccination with BNT162b2, Ad26.COV2.S, ChadOX1 at full, half and quarter doses. Radar plot showing as geometric mean titre (GMT) plasma neutralization of pseudovirus against Wild type (Wu-1), BA.1, BA.2 and XBB after two doses. GMT data is shown in log scale.

## Discussion

This study evaluated the safety and immunogenicity of fractional doses of three vaccines (ChadOx1, Ad26.COV2.S, and BNT162B2 vaccines) in Nigeria using a randomized triple-blinded non-inferiority clinical trial design. At 28 days post vaccination a total of 177 of 540 (21.7%) achieved seroconversion with the GMFR>2.5-fold rise in SARS-COV-2 antibody titre. In the ChadOx1 vaccine arm, the difference in seroconversion rates were -2.5% (95% CI: -19.9 to 15.0) for the half dose vs. full dose, and 3.2% (95% CI: -14.7 to 20.9) for the quarter vs Full dose and the in the Ad26.COV2.S vaccines the differences in the seroconversion were 5.7% (95% CI: -8.0 to 19.9) for the half dose vs. full dose and 2.9% (95% CI: -10.3 to 16.8) for the quarter vs. full dose; with both fractional doses considered non-inferior to the full doses. However, in the BNT162B2 vaccine arm, the fractional dose did not quite meet the non-inferiority criteria with a difference of -4.2% (95% CI: -20.5 to 12.1). No adverse events or severe adverse events were reported as the vaccines were safe and well-tolerated.

In this current study, the (quarter and half) fractional doses of ChadOx1 and Ad26.COV2.S vaccines induced adequate immune response among the participants similar to the full dose (standard dose) of the corresponding vaccines at day 28. The non-inferiority of the fractional doses to the full dose of the vaccines and the similarity in the participants who had adequate immunogenic response buttresses previous data coupled with predictive models of the efficacy of fractional doses of COVID-19 vaccines.^17^ Although the fractional dose (half) of BNT162B2 vaccines did not meet the non-inferiority criteria, GMFR a 2.5%-fold rise developed in similar proportions of participants (40% vs 44.4% for fractional and the full doses respectively). Furthermore, more participants seroconverted among the BNT162B2 group compared to those on either the fractional or full dose of ChadOx1 and Ad26.COV2.S vaccines.

In a subset of participants (32% of the study population) who were noted to be seronegative at baseline for anti-S IgG, it was noteworthy that 63% achieved seroconversion after receiving fractional vaccine doses across the three vaccines. The immunogenicity in this subpopulation was higher than seroconversion rates in the study population at 21.7%. The difference in seroconversion rates observed in the two groups may be attributed to a ceiling effect where antibody titres were near their peak due to repeated infections prior to vaccination. This finding is consistent with results from the UK and Israel, which described a ceiling or maximum S-RBD antibody titre level for antibody response. ^25–27^

In our sub-analysis of participants with longitudinal data across three-time points, vaccine-elicited neutralizing antibody and binding antibody responses were evaluated. We observed endemic level exposure to SARS CoV-2 characterized by positivity for IgG anti-N in 81% of participants at study entry and even higher levels for Omicron BA.1. This indicates extensive recent transmission of SARS CoV-2 virus before vaccination. The high degree of exposure is not surprising given that previous studies in the population observed 25% of SARS CoV-2 seroprevalence in 2020, almost 50% exposure rates in 2021^19,28^ and even higher rates were observed in a similar Nigerian population in 2023.^24^ Similar population-level studies in South Africa showed seroprevalence levels of 95% after the Omicron BA.1 wave.^29^ Additionally, IgG anti-N is expected to wane more rapidly than both anti-S and anti-RBD.^30^ When evaluating longitudinal response dynamics across the tested variants of concern, data showed broadening responses over time with neutralization titres, particularly for BA2, reflecting the impact of both vaccination and breakthrough infection in contributing to an enhanced and diversified immune repertoire which may not provide comprehensive protection as new variants emerge. Overall, the neutralization response data showed ≥2-fold response to both full and fractional doses indicating immunogenic responses of the vaccine dosing regimens and suggesting that the high degree of previous exposure provides similar rates of immune protection across the different dosing regimens.

The incidence of the reported adverse events in the trial was 9.1% and no severe adverse event was reported. This could be explained by the significant number of individuals who had developed immunity to the SARS-CoV-2 infection even though there was no previous history of infection and none of the participants had acute COVID-19 infection at enrolment. The incidence of adverse events reported is lower relative to trials in the region ^31,32,33^ and across other populations globally ^34,35,36^.. The plausible reason for the variation in the incidence could be a result of the adverse events reporting (active surveillance or self-report), individual immunogenetic differences, as well as gender, age, health status, time of the day that vaccine was received, and nutritional status.^37–40^

The common adverse events (irrespective of the vaccine arm or dose) were pain at the injection site, fever, tiredness, and body aches. These agree with reports by the World Health Organization and other previous studies in Nigeria^31,32,41^ and beyond^38,42–44^. The reported adverse event severity was mild to moderate in the current study with resolution of symptoms within 3 days post-vaccination. There was no report of life-threatening events such as rhabdomyolysis, inflammatory disorders associated with the heart (myocarditis or pericarditis), blood disorders (idiopathic thrombocytopaenia, thrombo-embolic disorders), anaphylaxis among others previously reported in some studies.^45–49^ In addition, the reported adverse events were self-limiting without the need for specialized care which aligns with earlier studies^48^. This affirms previous information and efforts towards correcting the misconception and myths of untoward adverse events associated with COVID-19 vaccines.

The pre vaccination seroprevalence among the study participants at 67.6% aligns with recent studies conducted in Nigeria. The seroprevalence reported by Akanmu et al. (60.3%) in Lagos^50^, 69.8% -80%, by Kolawole et al. in 12 states, including Lagos across the 6 geopolitical zones in Nigeria^51^ and 66.8% reported across six states by Olaleye et al^52^. Furthermore, similar reports (63%) by Sykes et al. among healthy populations in South Africa^53^, 56.8% in Somalia population by Hossain et al., 2023^54^ and a systematic review by Chisale et al.^55^

### Limitations

To our knowledge, this study is one of the first to report the safety and immunogenicity of fractional doses of primary course COVID-19 vaccines in sub-Saharan Africa, evaluating fractional doses of three approved COVID-19 vaccines used globally during the pandemic. While the trial also provides the advantage of having access to a large cohort of heterogeneous participants in a multi-site study such as this, the high attrition rate of the trial participants during the follow-up period significantly reduces the power to the generalizability of the outcome. This issue mirrors other studies that reported high attrition rates in Nigeria, Ghana and other sub-Saharan African countries on the completion of COVID-19 primary series vaccination.^19^ Another limitation was limited use of high-performance immunology assays such as the Luminex for IgG antibody measurement for the entire study population or an in-country prior performance evaluation validation of the selected assays given that assays can show different performance sensitivities in different populations.^56^

## Conclusion

The study showed that the fractional doses of ChadOx1 and Ad26.COV2.S vaccines were immunogenic and not inferior to their corresponding standard doses; it has potential to generate non-inferior immune responses compared to standard doses in the context of a population with high rate of previous exposure to SARS-CoV-2 infection. Neutralizing antibodies of all fractional vaccine types-ChadOx1, Ad26.COV2.S and BNT1-62B2 had good immunogenic responses against SARsCov-2 variants Overall, provides strong evidence for the feasibility of fractional dosing as a strategy to extend vaccine coverage in LMICs, with comparable immunogenicity to full dosing.

## Data Availability

All data produced in the present study are available upon reasonable request to the authors

## Acknowledgement

We extend our sincere appreciation to the trial participants in the study who repose their trust in the trial staff at the trial sites across the 5 geopolitical zones. Our depth of gratitude goes to the nurses, members of the data unit, and the counsellors/social workers of Aminu Kano University Teaching Hospital, Delta State University Teaching Hospital, Nnamdi Azikiwe University Teaching Hospital, Clinical Unit of National Institute for Pharmaceutical Research, Innovation and Development, the Clinical Trial Centre Nigerian Institute of Medical Research Trail who work tirelessly towards ensuring the successful outcome. Furthermore, our appreciation to the leaders and staff of the consortium institutions, the Nigeria Institute of Medical Research, National Primary Healthcare Development Agency, National Agency for Food Administration and Control and Nigerian Pharmaceutical Research, Innovation and Development

## Financial Support Information

Open Philanthropy

## Ethics declarations

All procedures performed in studies involving human participants were following the ethical standards of the institutional and/or national research committee and with the 1964 Helsinki Declaration and its later amendments or comparable ethical standards. This article does not contain any studies with animals performed by any of the authors.

## Informed Consent

Ethical approval was obtained from the National Health Research and Ethics Committee, the Institutional Review Board (IRB) of NIMR, Lagos, and the ethics committee of the trial institution. The purpose, processes, and expected outcome of the study were explained to all participants and/or their caregivers and informed consent was obtained before the commencement of the study. Confidentiality was maintained, and the freedom to withdraw at any time from the study without negative consequences was emphasized

---

**Supplementary Table 1.** Characteristics of COVID-19 Vaccines used for the study.

| Type of Vaccine | Doses/Vial | Composition | Full dose | Lot Number | Expiry Date |
| --- | --- | --- | --- | --- | --- |
| BNT162B2 | 6 | 30µg/dose | 0.3mL | FM7379 | 08/2022 |
| ChadOx1 | 10 | 5 x 10<sup>10</sup> vp/mL | 0.5mL | 4121Z260 | 07/2022 |
| Ad26.COV2.S | 5 | 1.0 x 10<sup>11</sup>vp/mL | 0.5mL | ACC5333 | 10/2023 |

**Supplementary Table 2.**
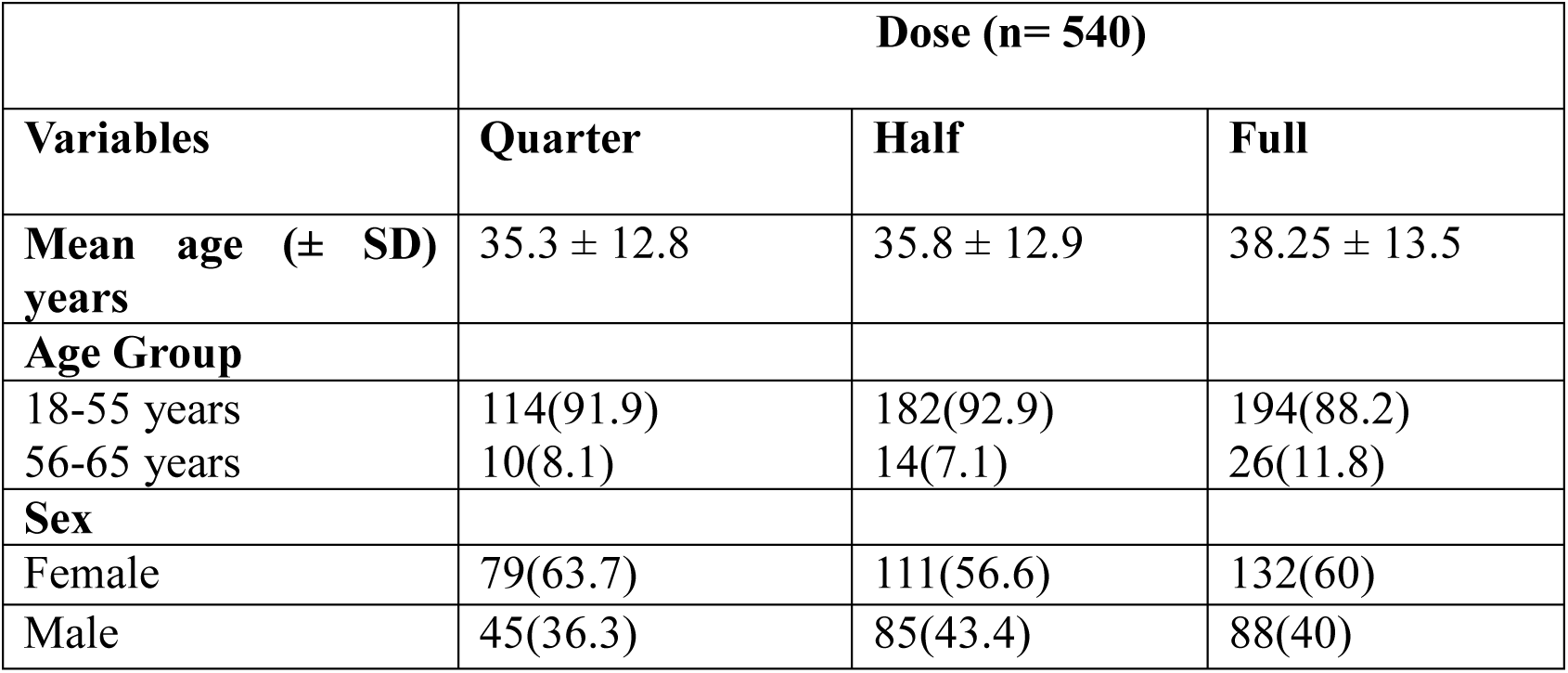
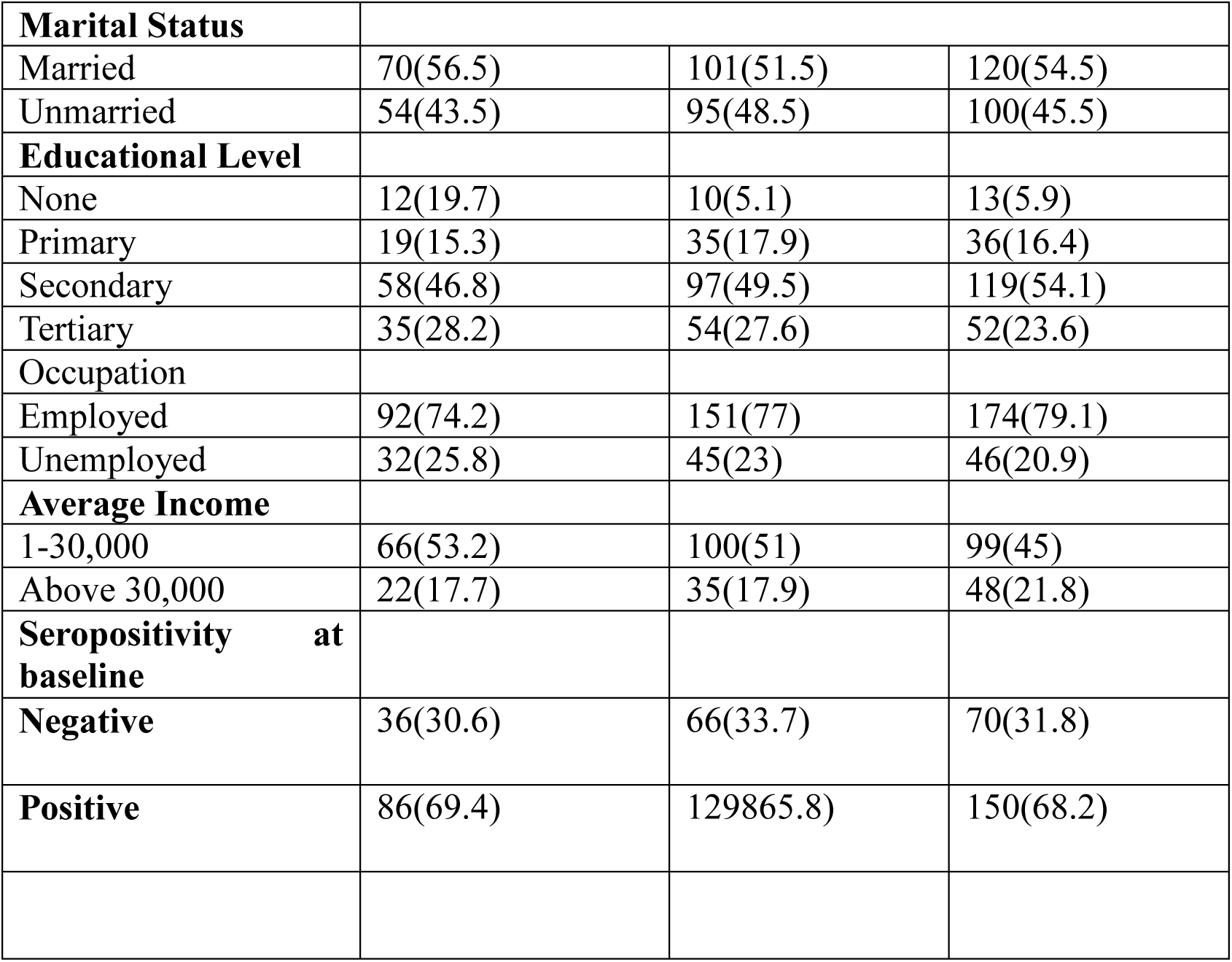
Baseline Characteristics of the Study Population based on the Primary Objective based on Vaccine dose (n= 540)

**Supplementary Table 3:** Adverse Events among the study participants by Vaccine type and Dose.

| Adverse events<br>(present) | ChadOx1 |  | BNT162B2 |  | Ad26.COV2.S |  |
| --- | --- | --- | --- | --- | --- | --- |
|  | Fractional doses<br>(Quarter/Half) | Full | Fractional<br>doses (Half) | Full | Fractional<br>doses<br>(Quarter/Half) | Full |
| Pain at injection site | 27 (43.5%) | 24 (38.7%) | 6 (16.7%) | 16 (44.4) | 22 (28.6%) | 32 (41.6) |
| Heavy arm | 2 (3.2%) | 0 (0.0%) | 0 (0.0%) | 2 (5.6%) | 0 (0.0%) | 1 (1.3%) |
| Itching at injection site | 0 (0.0%) | 4 (6.5%) | 0 (0.0%) | 0 (0.0%) | 1 (1.3%) | 0 (0.0%) |
| Swelling at injection<br>site | 0 (0.0%) | 0 (0.0%) | 1 (44.2%) | 0 (0.0%) | 0 (0.0%) | 0 (0.0%) |
| Rash at injection site | 0 (0.0%) | 0 (0.0%) | 0 (0.0%) | 0 (0.0%) | 0 (0.0%) | 0 (0.0%) |
| Fever | 5 (8.1%) | 9 (14.5%) | 5 (6.5%) | 7 (9.1%) | 2 (5.6%) | 4 (11.1%) |
| Chills and Rigor | 0 (0.0%) | 1 (1.6%) | 0 (0.0%) | 0 (0.0%) | 0 (0.0%) | 0 (0.0%) |
| Nausea | 0 (0.0%) | 0 (0.0%) | 0 (0.0%) | 0 (0.0%) | 0 (0.0%) | 0 (0.0%) |
| Body aches | 2 (3.2%) | 1 (1.6%) | 3 (8.3%) | 3 (8.3%) | 6 (7.8%) | 5 (6.5%) |
| Headaches | 2 (3.2%) | 4 (6.5%) | 3 (8.3%) | 2 (5.6%) | 2 (2.6%) | 2 (2.6%) |
| Insomnia (unable to<br>sleep) | 0 (0.0%) | 1 (1.6%) | 0 (0.0%) | 0 (0.0%) | 0 (0.0%) | 0 (0.0%) |
| Eye pain | 0 (0.0%) | 0 (0.0%) | 0 (0.0%) | 0 (0.0%) | 1 (1.3%) | 0 (0.0%) |
| Fatigue | 7 (11.3%) | 10 (16.1%) | 4 (11.1%) | 1 (2.8%) | 4 (5.2%) | 4 (5.2%) |
| Nasal discharge | 0 ( 0.0%) | 0 (0.0%) | 1 (2.8%) | 0 (0.0%) | 1 (1.3%) | 0 (0.0%) |
| Cough | 0 (0.0%) | 0 (0.0%) | 0 (0.0%) | 1 (2.8%) | 2 (2.6%) | 1 (1.3%) |
| Difficulty in breathing | 0 (0.0%) | 0 (0.0%) | 0 (0.0%) | 0 (0.0%) | 0 (0.0%) | 0 (0.0%) |
| Seizures | 0 (0.0%) | 0 (0.0%) | 0 (0.0%) | 0 (0.0%) | 0 (0.0%) | 0 (0.0%) |
| Loss of consciousness | 0 (0.0%) | 0 (0.0%) | 0 (0.0%) | 0 (0.0%) | 0 (0.0%) | 0 (0.0%) |
| Passage of watery stools | 0 (0.0%) | 0 (0.0%) | 0 (0.0%) | 0 (0.0%) | 0 (0.0%) | 0 (0.0%) |
| Vomiting | 0 (0.0%) | 0 (0.0%) | 0 (0.0%) | 0 (0.0%) | 1 (1.3%) | 0 (0.0%) |
| Redness of eye | 0 (0.0%) | 0 (0.0%) | 0 (0.0%) | 0 (0.0%) | 0 (0.0%) | 0 (0.0%) |
| Increased appetite | 1 (1.6%) | 0 (0.0%) | 1 (2.8%) | 1 (2.8%) | 1 (1.3%) | 1 (1.3%) |
| Slept a lot | 0 (0.0%) | 0 (0.0%) | 1 (2.8%) | 1 (2.8%) | 1 (1.3%) | 2 (2.6%) |

