## Supplementary material for "Safety and Immunogenicity of Fractional Doses of COVID-19 Vaccines among Nigerian Adults- A Randomised Non-Inferiority trial": supp

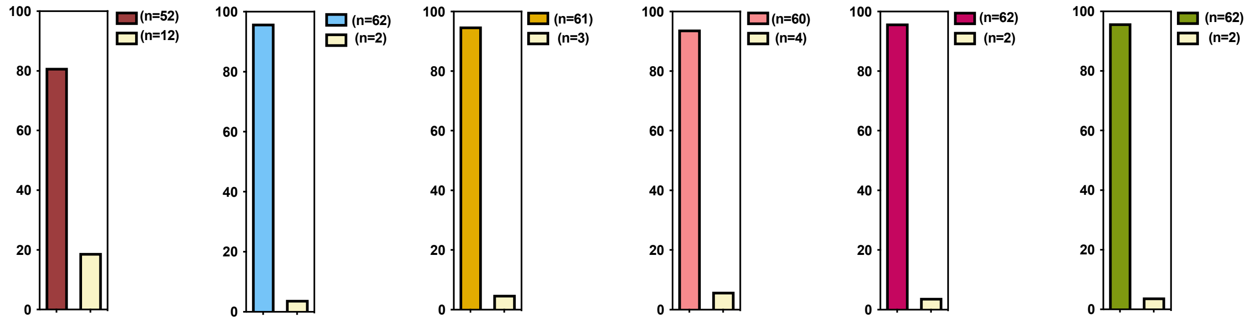


**Supplementary Figure 1: Luminex analysis of serum binding antibodies in a subset of participants**

Proportion of participants who were positive and negative for total IgG antibodies against SARS COV-2 Wu-1 anti-Nucleocapsid against SARS COV-2 Wu-1 anti-Nucleocapsid (anti-N), anti-Spike (S), anti-Receptor Binding Domain (Wu-1 RBD), anti-Receptor Binding Domain (Omicron BA.1 RBD), anti-Spike-1 (S1) and anti-Spike-1 Omicron (S1 Omicron) antibodies based on total number of participants recruited with samples available across three timepoints (n=64).

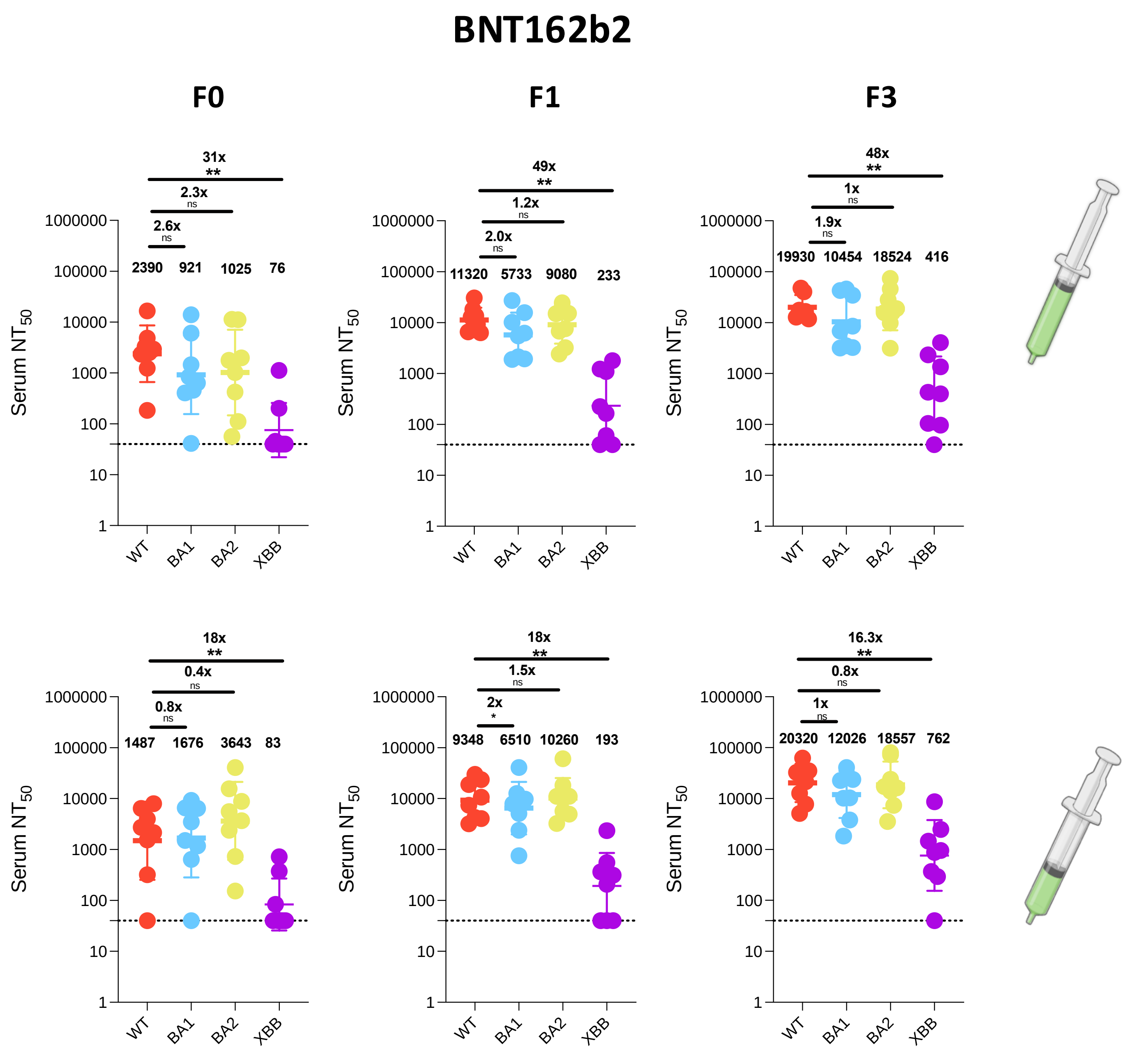


**Supplementary Figure 2: Antibody responses to vaccination with BNT162b2 at full and half doses**. a): Plasma neutralization of pseudovirus against Wild type (Wu-1), BA.1, BA.2 and XBB after two doses of the **BNT162b2** in Nigerian HIV-negative participants in Lagos Nigeria at three consecutive time points– baseline – F0 (before first-dose vaccination),F1 (1 month after 1st dose vaccination) and F5 (1-month post second dose at full doses (n=8) and half doses (n=8).

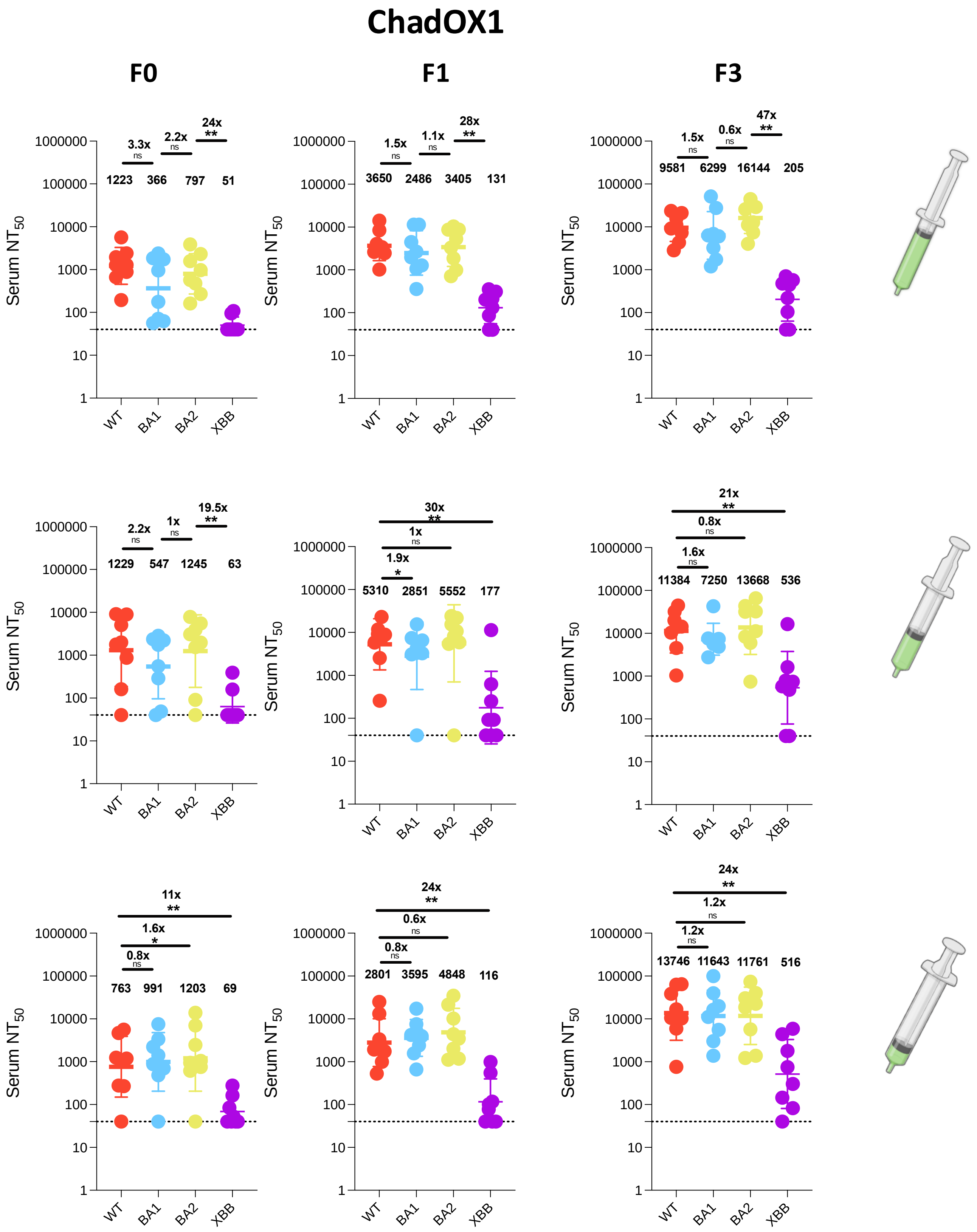


**Supplementary Figure 3: Antibody responses to vaccination with ChadOX1 at full, half and quarter doses.**

a): Plasma neutralization of pseudovirus against Wild type (Wu-1), BA.1, BA.2 and XBB after two doses of the **ChadOX1** in Nigerian HIV-negative participants in Lagos Nigeria at three consecutive timepoints– baseline – F0 (before first-dose vaccination), F1 (1 month after 1st dose vaccination) and F5(1-month post second dose at full, (n=8); half (n=8) and quarter doses (n=8).

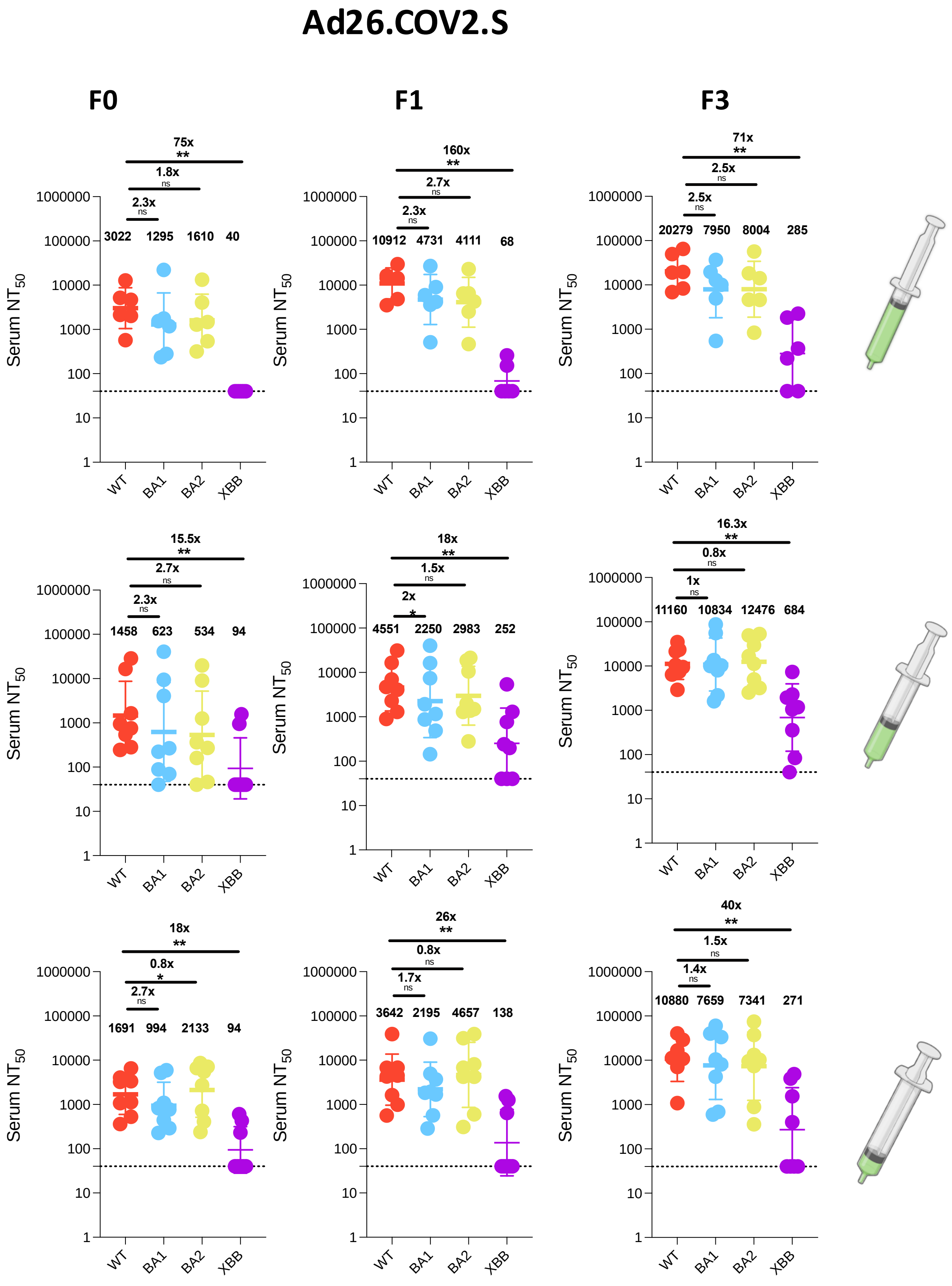


**Supplementary Figure 4 : Antibody responses to vaccination with Ad26.COV2.S at full, half and quarter doses.**

a): Plasma neutralization of pseudovirus against Wild type (Wu-1), BA.1, BA.2 and XBB after two doses of the Ad26.COV2.S in Nigerian HIV-negative participants in Lagos Nigeria at three consecutive time points– baseline – F0 (before first-dose vaccination), F1 (1 month after 1st dose vaccination) and F5 (1-month post second dose at full, (n=6); half (n=8) and quarter doses (n=8).
